# Evaluating the impact of school-based interventions on youth loneliness: A systematic review and meta-analysis

**DOI:** 10.64898/2026.04.15.26349177

**Authors:** Lou Sticpewich, Het Stuttard, Feifei Bu, Daisy Fancourt, Daniel Hayes

## Abstract

**Aims:** Youth loneliness is a prevalent global health concern with lifelong health ramifications. Schools, as children’s primary peer environments, are promising settings for loneliness interventions. However, school-based interventions are highly heterogeneous and no review to date has evaluated their effect on loneliness specifically.

**Methods:** A systematic review was conducted to identify studies of school-based interventions measuring loneliness as an outcome in children and young people aged up to 18. Meta-analyses were conducted using a random-effects model to pool effect sizes and examine the significance of intervention characteristics and study design. Reported implementation factors were extracted and narratively synthesised.

**Results:** Thirty-eight studies were included in meta-analysis, of which 19 were randomized controlled trials, ten were non-randomized controlled, and nine were single group studies. A small-to-moderate effect estimate was found, Hedges’ *g* = −0.42 [95% CI: −0.71, −0.13], *p* = .006, and sub-group analyses indicated that differences in study design and quality did not result in significantly different effect estimates. Psychological interventions, followed by social and emotional skills training, produced significantly higher effects estimates compared with other intervention types.

**Conclusions:** Findings indicate that school-based interventions are effective in reducing youth loneliness. However, study heterogeneity, reporting inconsistencies, and a wide prediction interval indicates this finding should be interpreted with caution. Future research may benefit from improved measurement and reporting of implementation factors, particularly dosage and fidelity.

## Introduction

Loneliness is highly prevalent among young people.^1,2^ Chronic loneliness experience in childhood and adolescence is associated with poorer health outcomes throughout the lifespan, from mental health issues to cardiovascular disease.^3–5^ Schools are crucial sites for understanding loneliness and delivering related interventions, as children and young people’s primary peer environment. Relatively few loneliness-specific interventions have been implemented in schools to date, but the wide array of interventions to promote social skills and positive interpersonal relationships within schools may implicitly influence young people’s loneliness experience.

Previous systematic reviews have synthesised the evidence base for youth loneliness interventions across settings or for school-based wellbeing interventions more broadly, but none has evaluated the impact of school-based interventions on loneliness specifically.^6,7^ Youth loneliness interventions are promising: previous meta-analysis found they significantly reduced loneliness with a moderate effect size across setting, intervention type, delivery mode, participant gender and age.^6^ These findings have limited utility to researchers and professional seeking to develop and assess effective interventions in schools to address loneliness rates. This new review focuses on children and young people across early and mandatory education settings to examine the impact of school-based interventions on youth loneliness and what factors influence their suitability and effectiveness.

## Methods

### Database Search

The review was prospectively registered on PROSPERO. Seven electronic databases were selected (MEDLINE, Embase, PsycINFO, Global Health, Social Policy and Practice, Scopus, and Web of Science Core Collection) and no search date restrictions applied. Three search topics were used – ‘youth,’ ‘loneliness,’ and ‘school-based interventions.’ ‘Loneliness’ included subjective and objective social deficit terms. ‘School-based intervention’ terms were permutations of ‘school’ terms, such as *teach, student*, or *lesson*, combined using adjacency operators to ‘intervention’ terms, like *referral, therapy*, or *prevention* (full strategy in **S1**). Title and abstract searches were run on 1 July 2025.

### Screening and Selection

Database searches returned 8,226 records. Results were extracted into EndNote and 3,350 duplicate records removed. The remaining records were imported into Rayyan, where an additional 14 duplicate records were removed. Studies were screened against the following criteria: (1) published in English; (2) sample mean age below 18 years old; (3) examined an intervention delivered, at least in part, in a school setting; and (4) measured loneliness before and after intervention.

Eligible studies did not require a control group. ‘School’ spanned early childhood education through compulsory education, across general and alternative education settings. Case studies, conference abstracts, and cross-sectional studies were excluded. Digital interventions and interventions in community, health, and home settings were eligible if school involvement was also specified, e.g. point of referral from staff, delivery during school hours, or on school premises. Where no abstract was available or information was missing, the study was progressed to full-text screening.

A total of 4,862 title and abstract records were screened for relevance. 488 records (10%) were double-screened, with 93% agreement and all conflicts resolved through discussion. 817 studies progressed to full text screening, while relevant reviews were separated out for reference searching. 82 full text studies (10%) were double-screened, with 91% agreement and seven conflicts resolved through discussion, resulting in 38 included studies: nine single group studies, ten non-randomised controlled studies and 19 randomised controlled trials (RCTs).

### Data Coding

Author, publication date, study design, participant characteristics (e.g. sample size, gender, age), intervention characteristics (e.g. type, duration, school pathway details, use of technology), loneliness measurement, and implementation factors were extracted from each included study. Authors were contacted regarding missing data and study design clarification. Study quality was assessed using the Effective Public Health Practice Project (EPHPP) Quality Assessment Tool.^8^

### Meta-analysis

Standardised mean differences (SMD) were calculated for each included study (*k =* 38) in R, using the mean loneliness change-from-baseline scores divided by the pooled standard deviation at baseline, to minimise error due to group differences at baseline and within-group intervention effects. Where studies included multiple follow-up measurements, only the first post-intervention loneliness score was used to calculate the SMD. For single group studies, a counterfactual matched group of the same size and standard deviation with a null mean change-from-baseline was assumed.^7^ For studies with multiple intervention groups, mean loneliness scores were combined and standard deviations pooled for comparison with the control group, to avoid including correlated effect sizes in the meta-analysis or double counting control groups. All SMDs were bias corrected, due to several studies with small sample sizes, to produce Hedges’ *g*.^9^ Effects sizes were pooled in R using the {*meta}* package, assuming a random-effects model. Heterogeneity was assessed using the *Q*-statistic, *τ* ^*2*^, and *I*^*2*^.*τ*^*2*^ was estimated using the restricted maximum likelihood method,^10^ with Knapp-Hartung adjustments to confidence interval calculations.^11^ Outliers were identified and removed to examine changes to heterogeneity statistics and the prediction interval of the effect estimate. Sub-group analysis was performed for study design and quality, intervention type and format, and school type as a rough proxy for age group.

## Results

Thirty-eight studies published between 1993 and 2025 were included in the random-effects meta-analysis. Study sample sizes varied from 10 to 5,219 participants, producing a total of 16,698 individuals (7,779 female; 46.6%). All participants were between 5 to 20 years of age, with sample means below 18. Fourteen studies were conducted in North America (37%), ten in Europe, eight in West Asia, three in Australia, and three in East Asia. Study and sample characteristics, along with effect sizes, are presented in Tables 1, 2 and 3. Twenty-three studies involved a universal intervention with no screening or diagnostic criteria, five studies targeted groups of children and young people indicated to be ‘at risk,’ e.g. due to reported bullying experience or elevated scores on screening instruments, and ten studies were specific to children and young people with diagnosed health conditions and/or receiving clinical support.

**Table 1.** Single group/uncontrolled study information (k = 9)

| Authors (date) | Intervention type | Intervention length | Measurement points | Age range (mean, SD) | Sample size<sup>a</sup> | Sample description | Primary outcome measured | Loneliness measure | Quality rating | Impact on loneliness | Effect size [95% CI] |
| --- | --- | --- | --- | --- | --- | --- | --- | --- | --- | --- | --- |
| Abu-Rasain (1998)<sup>12</sup> | Peer support | Ad hoc over 6 months (1-5 sessions) | 4: baseline, 1, 2, and 6 months | 15-20 (17.288, NR)<sup>b</sup> | 278 (0, 0%) | Students at an all-boys' school in Saudi Arabia | Loneliness | UCLA (20-item; Arabic) | Weak | $F(3) = 6.1890, p = .1028$ (not sig. across all 4 time points) | -0.12 [-0.28, 0.05] |
| Bostick and Anderson (2009)<sup>13</sup> | Social and emotional skills | 10 sessions (frequency and duration not specified) | 2: baseline and post | 8-9<sup>c</sup> (NR, NR) | 49 (25, 51%) | Public school students in North Carolina, US with high loneliness and social anxiety | Emotional adjustment (loneliness and social anxiety) | LSDQ (8-item; English) | Weak | $t = -4.95, p < .001$ (sig decrease at post-test) | -0.73 [-1.13, -0.32] |
| Bradley (2016)<sup>14</sup> | Peer support | Fortnightly (6 months) | 2: baseline and post | 11-12 (11.67, 0.49<sup>d</sup>) | 12 (4, 33.3%) | Autistic students from five urban, mainstream schools in England | Self-esteem, social satisfaction, and bullying | LSDQ (24-item; English) | Weak | $t = -8.52, p < .01$ (sig. decrease at post-test) | -1.24 [-2.13, -0.35] |
| Fox et al. (2024)<sup>15</sup> | Social and emotional skills | 30 mins weekly (12 weeks) | 2: baseline and 2 weeks post | 11-14 (13.15, 1.00) | 10 (2, 80%<sup>d</sup>) | Students in a special education school in a large, urban US city | Loneliness | UCLA (8-item; English) | Weak | $t = 3.94, p < .01$ (sig. decrease at post-test) | -0.52 [-1.41, 0.38] |
| Gilham et al. (2023)<sup>16</sup> | Other (GuysWork – social norms) | 60 mins weekly (10 weeks) | 2: baseline and post<sup>e</sup> | 12-13 (NR, NR) | 152 (0, 0%) | Male students at 14 schools in Nova Scotia, Canada, selected by school staff based on troubling social and emotional behaviours | Loneliness, perceived stress, and gender role norms | UCLA (6-item; English) | Moderate | $z = -0.76, p > .017$ (not sig.) | -0.12 [-0.37, 0.13] |
| King et al. (1997)<sup>17</sup> | Social skills (Joining In) | 2 x 90 mins weekly (10 weeks) | 3: baseline, 10 weeks, and 6 months | 8.6-14.6 (12.0, NR) | 11 (8, 72.7%) | Children with a diagnosis of cerebral palsy or spina bifida in Ontario, Canada | Social self-perception | LSDQ (24-item; English) | Weak | Not sig. at post-test, but sig. ($t = 2.73, p < .05$) at follow-up | -0.17 [-1.01, 0.67] |
| Kishida et al. (2022)<sup>18</sup> | Social and emotional support (web resource) | 6 x 40-60 min (10 weeks) | 2: baseline and 10 weeks | 10-12 (NR, NR) | 19 (10, 52.6%) | Deaf and hard-of-hearing students in mainstream schools in Western Australia | Social and emotional wellbeing | Loneliness at School Scale (7-item; English) | Weak | $t = -0.28, p = .782$ (no sig. pre-post difference) | 0.04 [-0.63, 0.71] |
| Kopelman-Rubin et al. (2012)<sup>19</sup> | Psychological intervention (I Can Succeed) | 13 sessions (3 months)<sup>f</sup> | 2: baseline and post | 11-15 (12.60, 0.87) | 40 (12, 30%) | Students diagnosed with learning disorders at junior high school in central Israel | Academic and emotional functioning | PNDLS (16-item; Hebrew) | Weak | No sig. change at post-test | -0.15 [-0.61, 0.31] |
| Margalit (1995)<sup>20</sup> | Social and emotional skills (I Found a Solution) | 2 x 45 mins weekly (4 months) | 2: baseline and 4 months | 11.0-15.0 (12.39, 1.24)<sup>g</sup> | 83 (0, 0%) | Male students with learning disabilities or behavioural difficulties in special education classes in Israel | Social skills | LSDQ (24-item; Hebrew) | Weak | $F(1, 77) = 4.54, p < .05$ (sig. pre-post comparison) | -0.28 [-0.59, 0.02] |

Note. NR = not reported. Negative direction of effect indicates decrease in loneliness.
<sup>a</sup> Sample size (number of female participants, % female participants)
<sup>b</sup> Age range and mean reported only for peer client sub-group ( $n = 66$ ) rather than whole sample, but considered to be representative as peer clients spread across all 3 included grades
<sup>c</sup> Age range not reported and estimated based on included school grades
<sup>d</sup> Mean age and SD calculated from raw ages of participants
<sup>e</sup> Additional follow-up planned but not analysed due to very low completion related to COVID pandemic
<sup>f</sup> Additional follow-up planned but not analysed due to very low completion related to COVID pandemic
<sup>g</sup> Additional follow-up planned but not analysed due to very low completion related to COVID pandemic
<sup>f</sup> 6 booster sessions offered over subsequent 18 months but effect not detectable as post-assessment completed at 3 months
<sup>g</sup> Mean age and SD reported for sub-groups – combined mean and SD calculated by first author

**Table 2.** Non-randomised controlled study information (k = 10)

| Authors (date) | Intervention type | Intervention length | Measurement points | Age range (mean, SD) | Sample size <sup>a</sup> | Sample description | Primary outcome measured | Loneliness measure | Quality rating | Impact on loneliness | Effect size [95% CI] |
| --- | --- | --- | --- | --- | --- | --- | --- | --- | --- | --- | --- |
| Allen-Kosál (2008)<sup>21</sup> | Other (cooperative learning) | 6 sessions pre-training and/or 2-4 x weekly activities (6-8 weeks) | 3: baseline, post, and 1 month post | 8-9<sup>b</sup> (NR, NR) | 72 (NR, NR) | Students from four elementary schools in the same county in Tennessee, US | Loneliness | LSDQ (24-item; English) | Weak | $F(6, 49) = 1.08, p = .39$ (non-sig. group x loneliness x time interaction, no sig. main effects) | 0.19 [-0.27, 0.65] |
| Gökcan and Yazici (2022)<sup>22</sup> | Social and emotional skills (play education program) | 90 mins weekly (11 weeks) | 2: baseline and post | 5-6 (5.67, NR) | 38 (NR, NR) | Kindergarten students in Kütahya city centre, Turkey | Loneliness | LSDQ (23-item; Turkish) | Moderate | $t = -5.409, p < .05$ (sig. difference between groups at post-test) | -3.73 [-4.82, -2.64] |
| Guthrie (1999)<sup>23</sup> | Other (classroom rule) | Duration of spring semester (approx. 17 weeks) | 2: baseline and end of school year (approx. 5 months) | 5-6 (NR, NR) | 92 (49, 53.2%) | Kindergarten students from seven public elementary schools in US Midwest | Peer acceptance, self-competence, loneliness, aggression | LSDQ (24-item; English) | Weak | $F = 1.06, p > .05$ (non-sig. difference between groups at post-test) | -0.20 [-0.63, 0.22] |
| Harrist and Bradley (2003)<sup>24</sup> | Other (classroom rule) | 30 mins weekly (15 weeks) | 2: baseline and end of school year | 5-6<sup>b</sup> (NR, NR) | 144 (71, 49.3%) | Kindergarten students attending three diverse elementary schools in Texas, US | Social acceptance | LSDQ (16-item; English) | Weak | $F = 5.44, p < .02$ (sig. main effect of group, controlling for baseline differences – note loneliness *increased* in intervention group) | 0.31 [-0.06, 0.69] |
| Morin (2022)<sup>25</sup> | Social support (VIP Partnership) | 9 weeks | 3: baseline, 10 weeks, and 6 months | NR (NR, NR) | 1835 (669, 61%) | Students at 17 secondary schools in Norway | Happiness, depression, anxiety and loneliness | LSDQ (Norwegian) | Moderate | $p = .019$ for between group $t$-test at 6-month follow-up, but no sig. main effects at post-test or follow-up using adjusted means | 0.00 [-0.09, 0.09] |
| Orkibi et al. (2017)<sup>26</sup> | Psychological intervention (psychodrama group therapy) | 16-22 x 90 mins (over school year) | 2: baseline and post | 13-16 (14.5, 0.78) | 40 (15, 37.5%) | Students from three middle schools in a low socioeconomic area of Israel | Self-concept and loneliness | LSDQ (24-item; Hebrew) | Weak | $F = 4.57, p < .05$ (sig. time x group interaction) | -0.58 [-1.26, 0.09] |
| Rointan et al. (2021)<sup>27</sup> | Psychological intervention (group play and painting therapy) | 45 mins weekly (8 weeks) | 3: baseline, post, and 45 days post | 11-12 (11.20, 1.22) | 45 (45, 100%) | Female students with specific learning disabilities in Kermanshah, Iran | Social adjustment and loneliness | LSDQ (24-item; Persian) | Moderate | $F = 60.69, p < .01$ at post-test and $F = 47.08, p < .01$ at follow-up (sig. group x time interaction) | -3.03 [-3.94, -2.14] |
| Stavrou et al. (2024)<sup>28</sup> | Other (human rights education) | 45 mins weekly (12 weeks) | 3: baseline, post, and 4 | 9-12 (11.2, | 340 (166, | Students from seven primary schools in | Human rights knowledge | Adapted LSDQ (5-item; | Weak | $F(1.17, 185.54) = 67.61, p = .000$, (sig. | -0.49 [-0.71, |
|  |  |  | months | 0.9) | 49.3%) | Thessaloniki, Greece | and school adjustment | Greek) |  | group x time interaction, with decrease in intervention group and increase in control) | -0.27] |
| Vassilopoulos, Brouzos and Koutsianou (2018a)<sup>29</sup> | Social and emotional skills | 45 mins weekly (7 weeks) | 3: baseline, post, and 3 months post | 6-7<sup>b</sup> (6.4, 0.27) | 113 (46, 40.7%) | Students from five medium-sized, inner-city public schools in north-western Greece | School adjustment | LSDQ (24-item; Greek) | Weak | $F(1.77, 196.85) = 0.98, p = .368$ (non-sig. time x group interaction) | -0.19 [-0.56, 0.18] |
| Zhou and McLellan (2024)<sup>30</sup> | Social and emotional skills | 40 mins weekly + ad hoc meditation practice (8 weeks) | 1: baseline and post | 12-14 (13.03, 0.50) | 362 (172, 47.5%) | Students at an urban, private school in China | Social and emotional wellbeing | UCLA (20-item; Chinese) | Weak | $t = -2.37, p < .05$ for taught group pre-post change, $t = .33$ for self-help and $t = .41$ for control ($p > .05$), no group sig. diff. in linear mixed model ($p > .10$) | -0.09 [-0.31, 0.13] |

Note. NR = not reported. Negative direction of effect indicates decrease in loneliness.
<sup>a</sup> Sample size (number of female participants, % female participants)
<sup>b</sup> Age range not reported, estimated based on included school grades

**Table 3.**
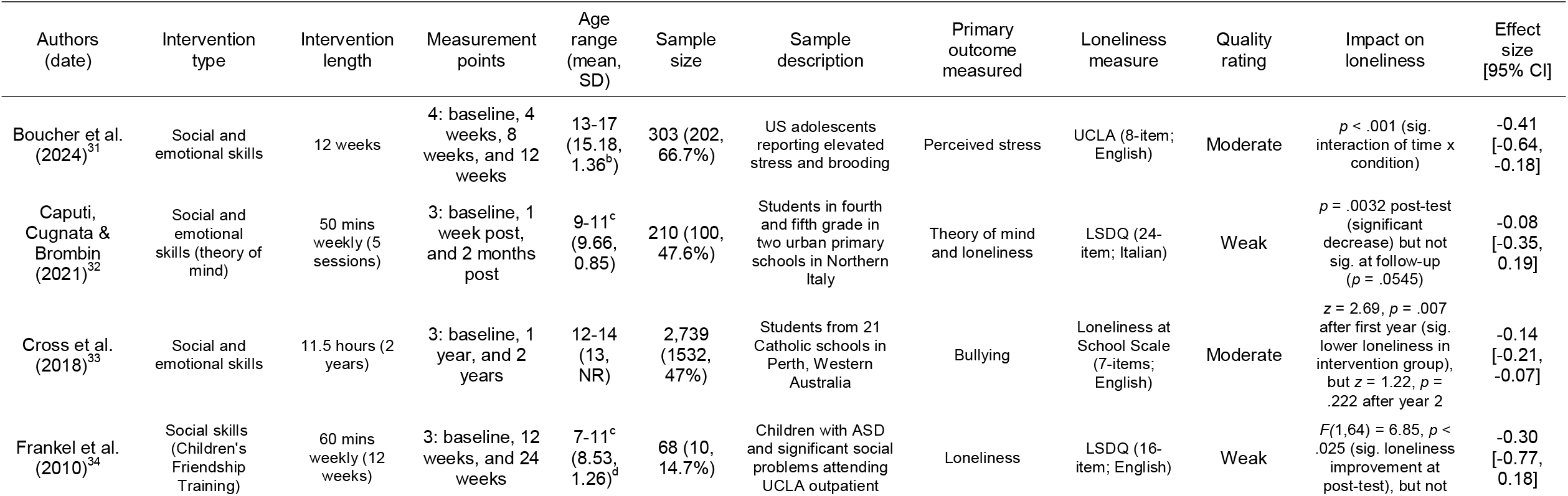
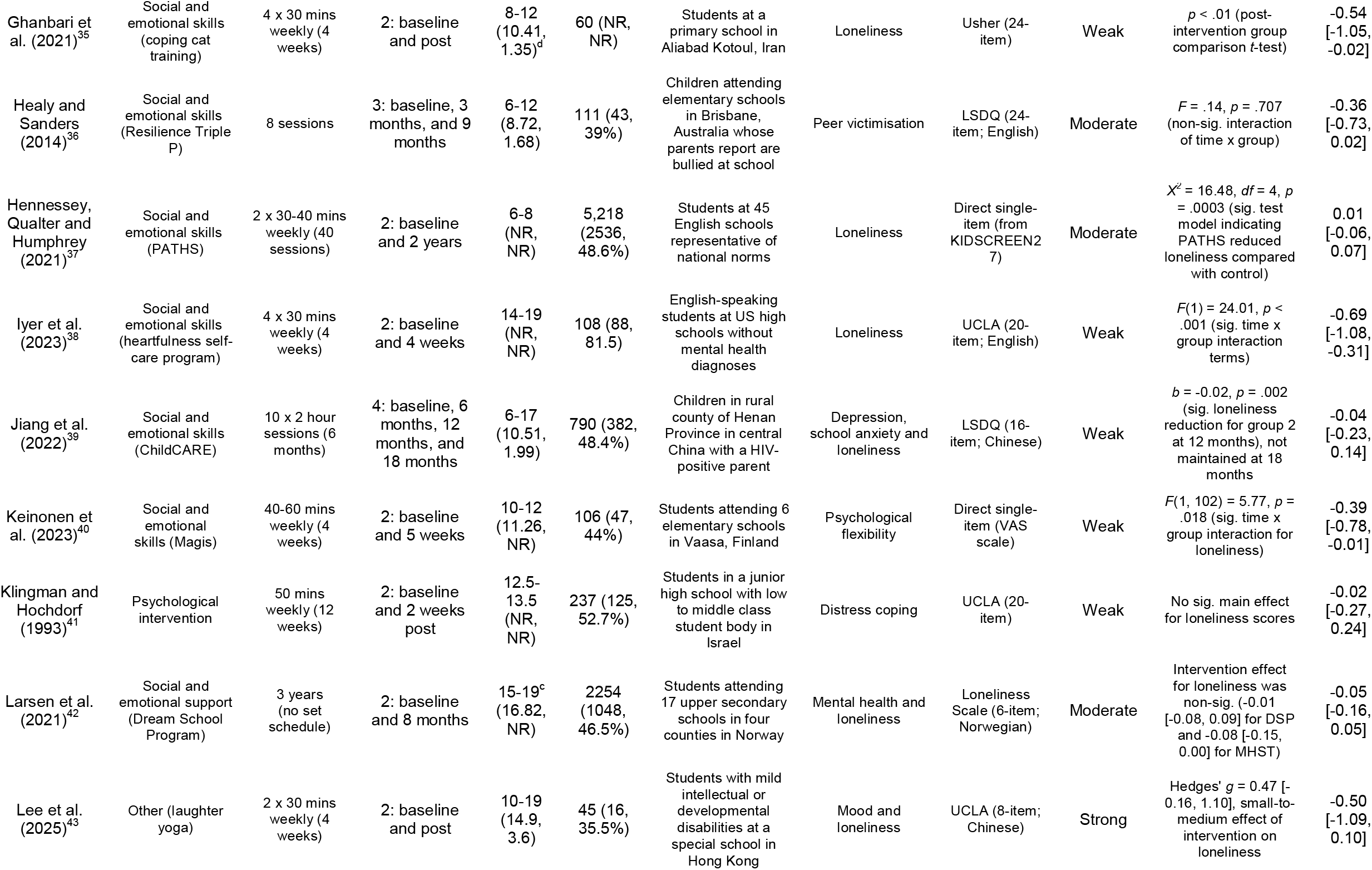
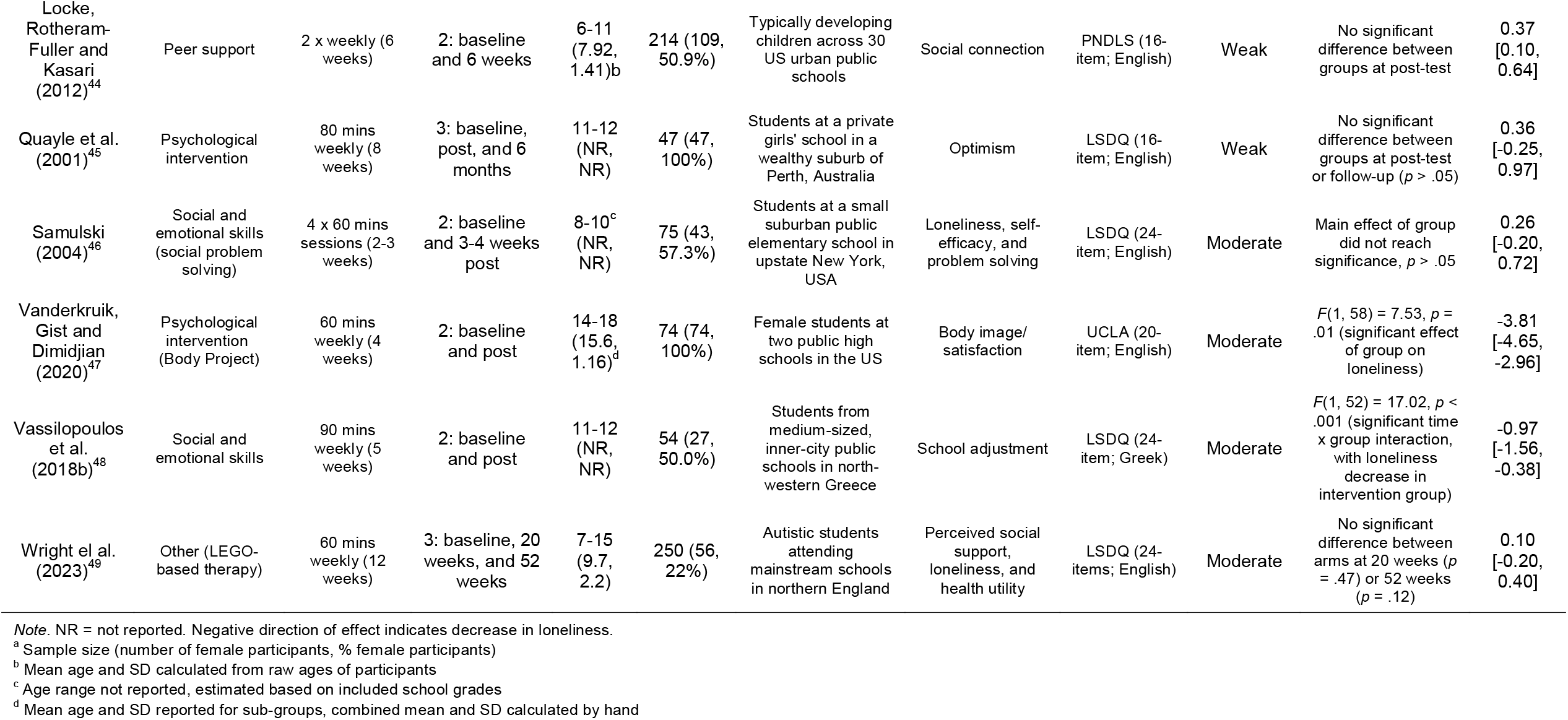
Randomised controlled study information (k = 19)

The majority of studies (*k =* 30) examined group interventions, with six studies of individual interventions and two of interventions with both group and individual components. Nineteen studies featured interventions predominantly comprised of social and emotional skills training, six involved a psychological intervention that was manualised and/or delivered by a trained professional, three enhanced levels of peer support (e.g. mentoring), two improved social and emotional support for young people in their schools, and eight were classed as ‘other.’ Three studies were conducted in kindergarten, 14 in primary schools, 18 in secondary schools, and three across primary and secondary school age groups. Five studies evaluated interventions that were entirely digital and two interventions with important digital components, but most studies (*k =* 31) were non-digital in format and delivery.

More than half of included studies (*k =* 24) were quality assessed as ‘weak.’ Thirteen studies were assessed as ‘moderate’, and only one as ‘strong’. Twenty-five studies (66%) were pilots. Detail provided on intervention delivery was varied and often limited, particularly regarding dosage and fidelity. School engagement in interventions varied significantly: in five studies, school involvement was limited to support with recruitment or outcome measurement. In sixteen studies, schools were the intervention setting, but staff were minimally involved with its implementation and delivery. Fifteen studies featured interventions that were in part or entirely delivered by school personnel, e.g. teachers, counsellors, or specialist staff, and two studies involved whole school interventions targeting school-wide systems alongside students and their caregivers.

The random-effects model estimated the pooled effect size of school-based interventions on loneliness to be *g* = −0.42 [95% CI: −0.71, −0.13], *p* = .006. The forest plots below (Figures 2, 3 and 4 – refer to **S2** for forest plot of all studies) depict the distribution of effect size across included studies, from *g* = −3.81 to 0.37. Negative effect estimates indicate a decrease in loneliness levels: where loneliness scales with an opposite direction were used, the sign of the mean difference was reversed. The between-study heterogeneity variance was estimated as *τ*^*2*^ = 0.60 [95% CI: 0.44, 1.38], with an *I*^*2*^ value of 86% [95% CI: 81.8%, 89.3%] suggesting substantial heterogeneity. While the pooled effect size is small-to-moderate and significant, it is tempered by a wide prediction interval, *g* = −2.00 to 1.17, including zero. Seven outliers with non-overlapping confidence intervals with the pooled effect estimate were identified and removed. The re-calculated random-effects model for the remaining 26 studies had a smaller significant effect estimate of *g* = −0.23 [-0.34, −0.12], *p* = .0002, with *τ*^*2*^ = 0.04 [0.02, 0.16], with an *I*^*2*^ value of 62% [43.5%, 74.5%], indicating moderate heterogeneity persisted despite the removal of outliers. Funnel plot and Egger’s test indicate significant asymmetry (β = −1.96, SE = −0.59, t = −3.44, p = .002) due to small-study effects and between-study heterogeneity (see **S3**).

**Figure 1.**
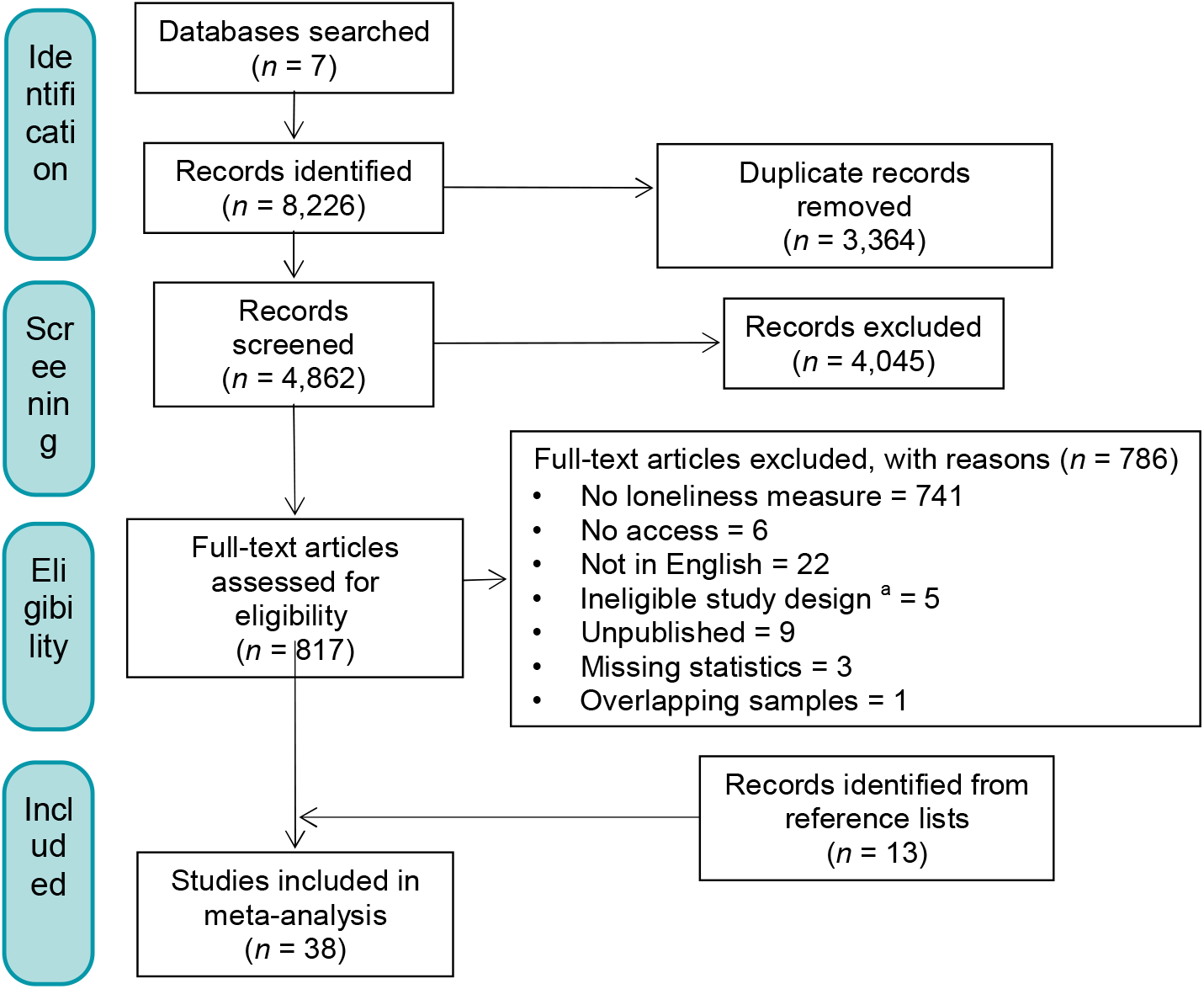
PRISMA flow diagram of study inclusion *Note*. ^a^ Ineligible study design included solely qualitative methods and studies without both pre- and post-intervention measurement of loneliness

**Figure 2.**
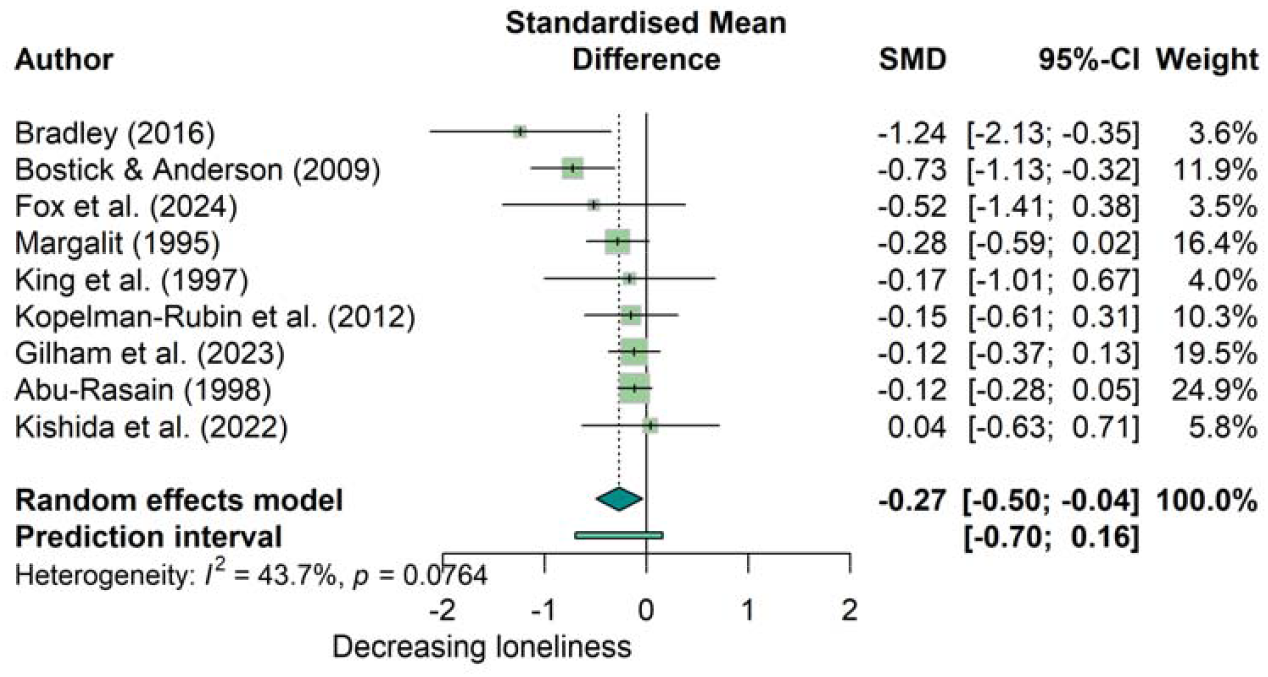
Forest plot of effect sizes for single group studies (k = 9)

**Figure 3.**
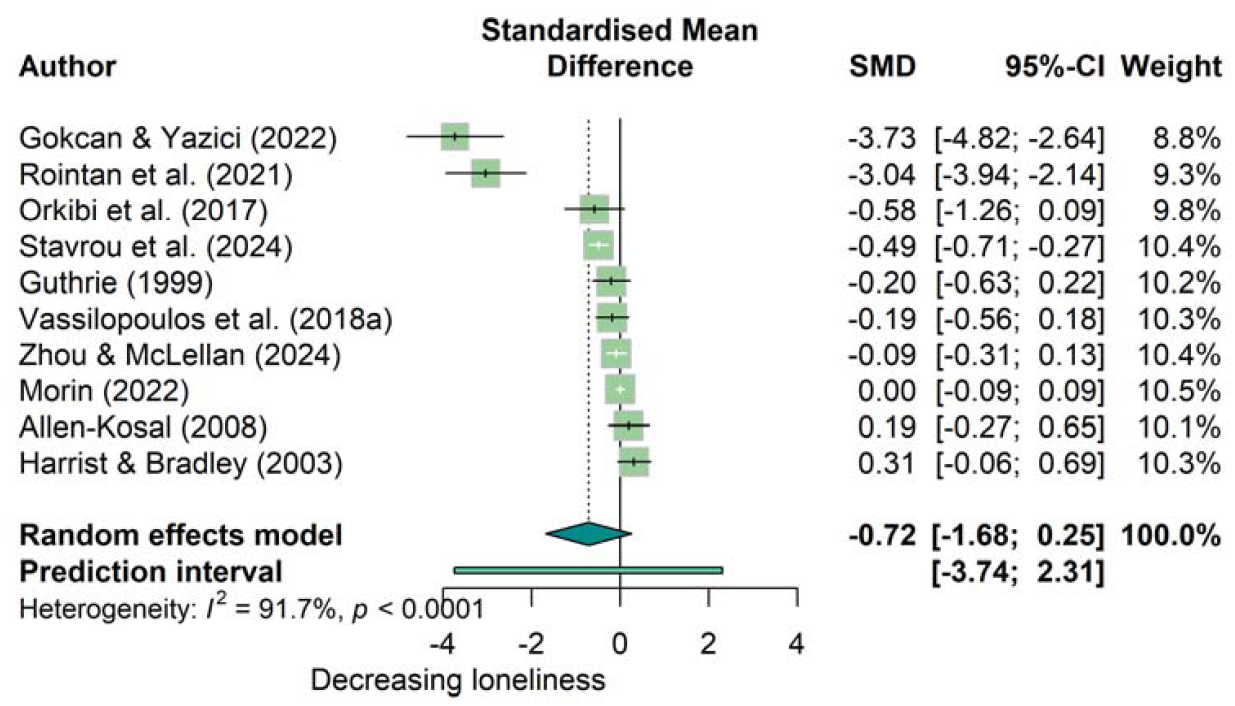
Forest plot of effect sizes for non-randomised controlled studies (k = 10)

**Figure 4.**
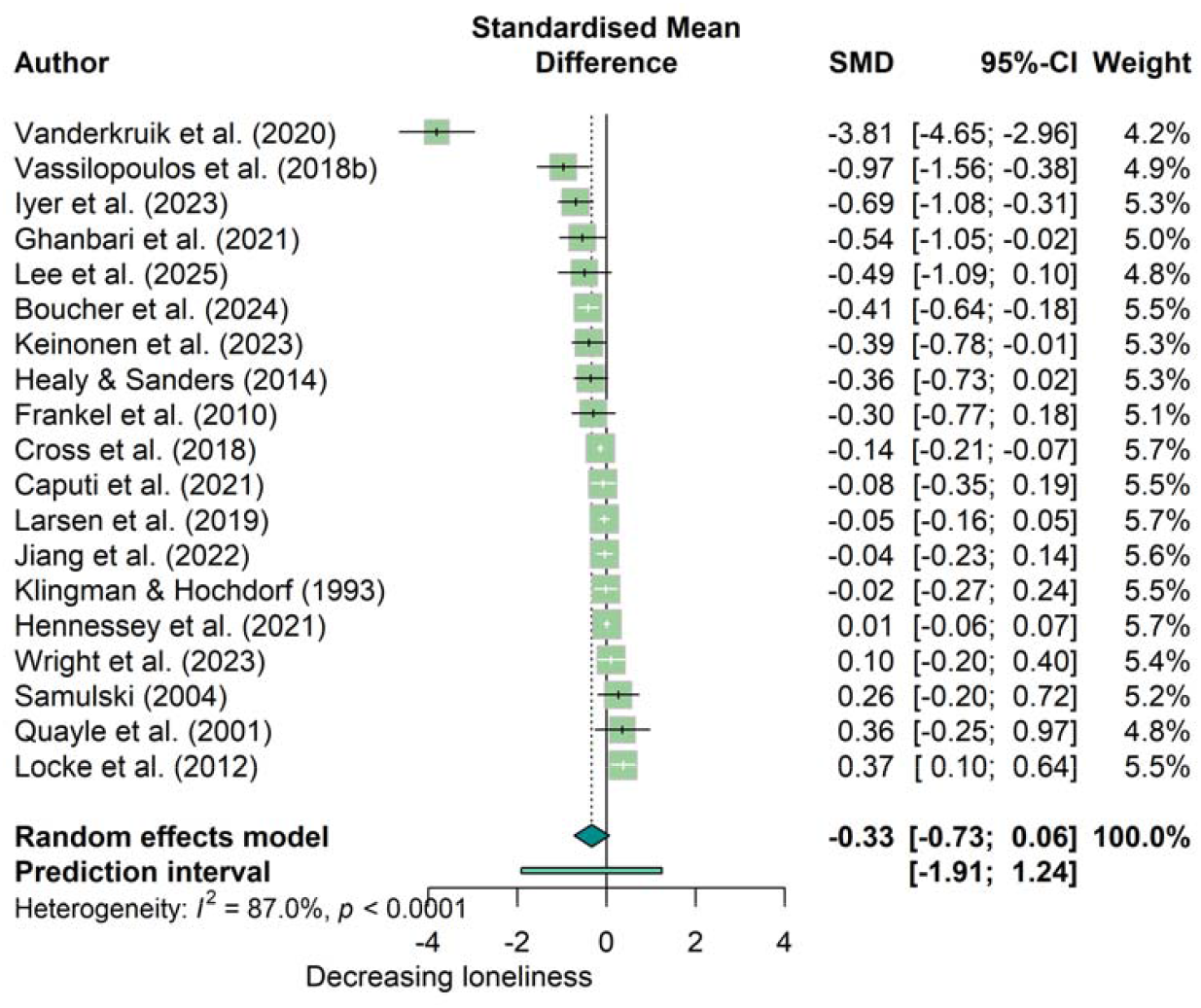
Forest plot of effect sizes for randomised controlled studies (k = 19)

### Subgroup Analyses

Subgroup analysis by study design was not significant (*Q* = 1.07, *df* = 2, *p* = .58). The largest pooled effect estimate and highest heterogeneity was seen in non-randomised controlled studies (*k* = 10, *g* = −0.72 [95% CI: −1.68, 0.25], *I*^*2*^ = 91.7%), followed by RCTs (*k* = 19, *g* = −0.33 [-0.73, 0.06], *I*^*2*^ = 87.0%) and single group studies (*k* = 9, *g* = −0.27 [-0.50, −0.04], *I*^*2*^ = 43.7%). Grouping by study quality was also not significant (*Q* = 3.66, *df* = 2, *p* = .16). Studies assessed as ‘moderate’ were more heterogeneous with a larger effect estimate (*k* = 13, *g* = −0.88 [-1.77, 0.01], *I*^*2*^ = 93.8%) than ‘weak’ studies (*k* = 24, *g* = −0.19 [-0.33, −0.05], *I*^*2*^ = 65.0%). Subgroup analysis by intervention type was significant (*Q* = 18.64, *df* = 6, *p* = .005), with psychological interventions (*k =* 6) producing the largest effect estimate of *g* = −1.18 [-3.01, 0.66] with substantial heterogeneity, *I*^*2*^ = 95.5%. Social and emotional skills training had the second largest effect estimate (*k* = 17, *g* = −0.41 [-0.77, −0.05], *I*^*2*^ = 84.2%), followed by peer support (*k* = 3, *g* = −0.24 [-2.18, 1.70], *I*^*2*^ = 88.0%). In contrast, social and emotional support (*k* = 1) and social support (*k* = 1) both had positive effect estimates of *g* = 0.04 [-0.63, 0.71] and *g* = 0.00 [-0.09, 0.09] respectively.

Intervention format was significant in subgroup analysis (*Q* = 15.41, *df* = 2, *p* = .0005). Group delivery (*k* = 30) had an effect estimate of *g* = −0.47 [-0.85, −0.09], *I*^*2*^ = 88.1%, individual delivery (*k* = 6) of *g* = −0.35 [-0.58, −0.11], *I*^*2*^ = 53.3%, and combined delivery of *g* = −0.05 [-0.22, −0.13]. Use of technology was also significant (*Q* = 10.76, *df* = 2, *p* = .005), with similar effect sizes for both non-digital and digital delivery (*k* = 31, *g* = −0.45 [-0.81, −0.08] and *k* = 5, *g* = −0.42 [-0.60, −0.25]).

Combined digital and non-digital delivery (*k* = 2) had a small effect estimate with *g* = −0.04 [-1.33, −1.24]. Grouping by school type was marginally significant, with *Q* = 8,58, *df* = 3, *p* = .035. Kindergarten (*k* = 3) had both the largest effect size and highest *I*^*2*^-statistic (*g* = −1.16 [-6.58, 4.26], *I*^*2*^ = 95.8%), followed by secondary (*k* = 18, *g* = −0.58 [-1.09, −0.07], *I*^*2*^ = 88.7%) and primary (*k* = 14, *g* = −0.21 [-0.42, −0.01], *I*^*2*^ = 78.9%). Delivery across primary and secondary school yielded the smallest effect estimate, *g* = −0.01 [-0.22, 0.20].

### Implementation Factors

Thirty-six studies discussed factors influencing intervention implementation and delivery, although with highly variable levels of detail (see **S4**). Competing pressures and unreasonable expectations faced by school staff were highlighted as implementation barriers,^13,46^ along with time, resource, and financial constraints faced by schools more broadly.^31,43,49^ Several studies suggested that short intervention durations may reduce impact and suggested longer, more intensive, embedded approaches.^25,32,36,41,42,46^ Students also faced high levels of demand that made participation challenging, exacerbated by intervention sessions scheduled during break periods or clashing with other activities.^30,45,47^

Studies of digital interventions highlighted their non-intrusive, flexible, and inexpensive features as beneficial to improve accessibility and surmount the challenges of school-based delivery.^15,31,38^ Universal interventions were described as cost-effective alternatives to targeted early intervention, as they do not require additional resource to identify at-risk students.^25,40^ Two interventions were delivered during class time already allocated to wellbeing curricula, to minimise disruption and facilitate easy integration.^16,30^ Multiple studies emphasised that high level of engagement from school staff was a crucial facilitator of intervention success, due to teachers’ trusted position and ability to adapt intervention content to fit students’ individual needs.^16,19,25,28,29,48^ The corollary noted in several studies was lack of enthusiasm from teachers severely constrained intervention efficacy, reducing intervention fidelity or dosage.^12,21,46^ Pre-existing differences between intervention facilitators, e.g. in style, experience, and training, introduces an additional source of variance, which was not evaluated in any of the included studies.^23,28^

Several studies discussed differential effects specific to universal classroom interventions.^23,24,29,45^ Where students do not report elevated levels of loneliness on average, studies are unlikely to detect an effect, even if interventions are affecting students with high social status and connectedness differently to students experiencing bullying or loneliness. For instance, Paley’s ‘You Can’t Say You Can’t Play’ rule appears to have increased social dissatisfaction among highly socially networked children, perhaps by limiting their ability to choose playmates.^24^ However, in-person group delivery within schools was described favourably by several studies as providing children and young people with opportunities to practice social skills and experience social satisfaction within naturalistic, socially valid environments.^15,17,22,23,41^

## Discussion

This systematic review and meta-analysis examined the effect of school-based interventions on loneliness in children and young people aged up to 18 years. The overall pooled effect estimate was *g* = −0.41 [95% CI: −0.67, −0.15], which was statistically significant (*p* = .003), indicating that school-based interventions can reduce loneliness experienced by children and young people. However, the wide prediction interval of this effect, *g* = −1.84 to 1.03, alongside large heterogeneity statistics (*I*^*2*^ = 86%), suggests substantial variation in the true effects necessitating cautious interpretation of the estimated effect. Very small or adverse effects may be possible in studies: one included study reported a significant increase in loneliness in the intervention arm,^24^ while another seven studies – varied in intervention type and target – reported smaller, non-significant increases.^18,21,37,44–46,49^ This is unsurprising given the diversity of included studies, which examined the effects of interventions with multiple components on varied samples of children and young people attending a range of kindergarten, primary, secondary, private, public, and special education schools across 14 different countries. Stratification by study quality or design did not meaningfully reduce between-study heterogeneity in subgroups or increase the precision of their effect estimates; few studies realised a ‘gold standard’ RCT design and many were pilot trials.

### Impact of Intervention Characteristics

Subgroup analysis by intervention type was significant and found that psychological interventions produced the largest effect estimate, followed by social and emotional skills training. This is in line with Lasgaard et al.’s review of loneliness interventions across the life-course, which included 84 psychological intervention studies and 52 social and emotional skills training studies.^7^ Grouping by intervention format and use of technology was significant, due to the small effect size and number of studies that used a combined group-individual format or digital with non-digital components.

Group delivery had a comparable effect estimate and 95% CI to individual delivery, although much more between-study heterogeneity, as did digital to non-digital delivery, which mirrors Eccles & Qualter’s finding that neither technology use nor delivery mode significantly moderated intervention effects.^6^ These findings corroborate the importance of balancing implementation factors with intervention aims: the intervention type and format matters insofar as it is appropriately matched to the intervention setting and population. Contending with a continuum of social connectedness within student groups is an important consideration for future research.

Subgroup analysis by school type found a marginally significant difference between groups, with interventions set in kindergartens demonstrating a large effect, medium effect in secondary schools, and small in primary schools, all with substantial between-study heterogeneity statistics. Included studies were close to evenly split between primary and secondary school settings – an improvement on Eccles and Qualter’s review, which only included six studies aimed at children.^6^ However, this finding was influenced by the decision to split kindergarten from primary school. If combined, the effect estimate would be close to that of the secondary school group. Wide groups risk masking distinct effects, but more precise classification is hindered by imprecision in study reporting and relatively few studies.

### Strengths and Limitations

Using a counterfactual null change control group to calculate the SMD of single group studies is likely to have over-estimated their effect sizes: control groups generally report some decrease in loneliness over time, in line with general trends over the school year. However, subgroup analysis by study design found single group had a smaller effect estimate than controlled studies, indicating that this over-estimation is not inflating the overall pooled effect beyond the effect estimated using measured controls. Differentiating between loneliness intervention approaches using meta-analytic techniques is fundamentally constrained by the diverse quality of existing research. Without improved methodological rigor and intervention reporting,^50^ meaningfully comparing or combining the effects of different interventions will continue to present difficulties. Given more information about data collection – for instance, whether children’s loneliness outcomes were verbally administered and self-rated, or collected by teachers, which was not stipulated in the majority of included studies – and implementation factors, particularly dosage and fidelity, more precise conclusions can be drawn about specific intervention suitability in school settings.

## Conclusions

This meta-analysis updates and synthesises the existing evidence base for loneliness interventions in children and young people by focusing on interventions delivered through education settings. In previous reviews of loneliness research, ‘children and young people’ included young adults up to 25 years old, meaning that studies specific to university students, already over-represented in psychology and behavioural science research, are folded into estimates of effect. While psychological interventions produced the largest effect estimate, the high level of between-study heterogeneity requires cautious interpretation of this finding. Rather than indicating a single ‘best’ school-based intervention, this review suggests that their success in tackling loneliness may have more to do with implementation context, such as school engagement, and finer-grained delivery factors like facilitator style and flexible scheduling, and these factors would benefit from further study.

## Ethical approval and informed consent statements

Not applicable for this study.

## Data availability statement

The raw data supporting the conclusions of this article will be made available by the corresponding author without undue reservation.

## Acknowledgements

We are grateful to Debora Marletta, UCL Library Services, for her support in refining the electronic search strategies. The author(s) declared no potential conflicts of interest with respect to the research, authorship, and/or publication of this article.

This research is funded by the Kavli Trust (Kavli2023-0000000064) and the Prudence Trust (INSPYRE, PT-0040). The funders do not play a role in study design, data collection, analysis, or reporting. Ethical approval was not applicable for this study. The raw data supporting the conclusions of this article will be made available by the corresponding author without undue reservation.

