## Supplementary Material for "Evaluating the impact of school-based interventions on youth loneliness: A systematic review and meta-analysis"

### S1. Full Search Syntax

In each database, search terms are within each search topic were connected by OR Boolean operators and the three search topics (youth, loneliness, and school-based interventions) are connected by AND operators.

|  | **Ovid** | | | | | **Web of Science core collection** | **Scopus** |
| --- | --- | --- | --- | --- | --- | --- | --- |
|  | **Medline** | **Embase** | **PsycInfo** | **Social Policy & Practice** | **Global Health** |  |  |
| Youth | (child* not childbirth).ti,ab,kf. | (child* not childbirth).ti,ab,kf. | (child* not childbirth).ti,ab,id. | (child* not childbirth).ti,ab. | (child* not childbirth).ti,ab. | TS=((child* not childbirth) or adolescen* or teenage* or "young person" or "young people" or youth) | (child* not childbirth) or adolescen* or teenage* or "young person" or "young people" or youth |
|  | adolescen*.ti,ab,kf. | adolescen*.ti,ab,kf. | adolescen*.ti,ab,id. | adolescen*.ti,ab. | adolescen*.ti,ab. |  |  |
|  | teenage*.ti,ab,kf. | teenage*.ti,ab,kf. | teenage*.ti,ab,id. | teenage*.ti,ab. | teenage*.ti,ab. |  |  |
|  | young person.ti,ab,kf. | young person.ti,ab,kf. | young person.ti,ab,id. | young person.ti,ab. | young person.ti,ab. |  |  |
|  | young people.ti,ab,kf. | young people.ti,ab,kf. | young people.ti,ab,id. | young people.ti,ab. | young people.ti,ab. |  |  |
|  | youth.ti,ab,kf. | youth.ti,ab,kf. | youth.ti,ab,id. | youth.ti,ab. | youth.ti,ab. |  |  |
|  | *Subject headings:*  exp Child/ or Adolescent/ | *Subject headings:*  exp child/ or exp adolescent/ | *Subject headings:*  - |  | *Subject headings:*  exp children/ or  adolescents/ |  |  |
| Loneliness | social isolation.ti,ab,kf. | social isolation.ti,ab,kf. | social isolation.ti,ab,id. | social isolation.ti,ab. | social isolation.ti,ab. | TS=("social isolation" or "social alienation" or "social deprivation" or "social detachment" or "social disconnect*" or victimi$ation or "social exclusion" or lonel*) | "social isolation" or "social alienation" or "social deprivation" or "social detachment" or "social disconnect*" or victimi?ation or "social exclusion" or lonel* |
|  | social alienation.ti,ab,kf. | social alienation.ti,ab,kf. | social alienation.ti,ab,id. | social alienation.ti,ab. | social alienation.ti,ab. |  |  |
|  | social deprivation.ti,ab,kf. | social deprivation.ti,ab,kf. | social deprivation.ti,ab,id. | social deprivation.ti,ab. | social deprivation.ti,ab. |  |  |
|  | social detachment.ti,ab,kf. | social detachment.ti,ab,kf. | social detachment.ti,ab,id. | social detachment.ti,ab. | social detachment.ti,ab. |  |  |
|  | social disconnect*.ti,ab,kf. | social disconnect*.ti,ab,kf. | social disconnect*.ti,ab,id. | social disconnect*.ti,ab. | social disconnect*.ti,ab. |  |  |
|  | victimi?ation.ti,ab,kf. | victimi?ation.ti,ab,kf. | victimi?ation.ti,ab,id. | victimi?ation.ti,ab. | victimi?ation.ti,ab. |  |  |
|  | social exclusion.ti,ab,kf. | social exclusion.ti,ab,kf. | social exclusion.ti,ab,id. | social exclusion.ti,ab. | social exclusion.ti,ab. |  |  |
|  | lonel*.ti,ab,kf. | lonel*.ti,ab,kf. | lonel*.ti,ab,id. | lonel*.ti,ab. | lonel*.ti,ab. |  |  |
|  | *Subject headings:*  exp Social Isolation/ | *Subject headings:*  social isolation/ or social alienation/ or loneliness/ | *Subject headings:*  Social Deprivation/ or Social Isolation/ or Loneliness/ or Social Exclusion/ |  | *Subject headings:*  social isolation/ or loneliness/ |  |  |
| School-based interventions | (teacher* adj3 referral*).ti,ab,kf. | (teacher* adj3 referral*).ti,ab,kf. | (teacher* adj3 referral*).ti,ab,id. | (teacher* adj3 referral*).ti,ab. | (teacher* adj3 referral*).ti,ab. | TS=((teacher* NEAR/2 referral*) or (school* NEAR/2 referral*) or (student* NEAR/2 referral*) or (pupil* NEAR/2 referral*) or (school* NEAR/2 intervention*) or (preschool* NEAR/2 intervention*) or (kindergarten* NEAR/2 intervention*) or (class* NEAR/2 intervention*) or "teach* intervention*" or "learning intervention*" or (student* NEAR/2 support) or (pupil* NEAR/2 support) or (school* NEAR/2 support) or (preschool* NEAR/2 support) or (kindergarten* NEAR/2 support) or (lesson* NEAR/2 support) or (class* NEAR/2 support) or (teach* NEAR/2 support) or (learning NEAR/2 support) or (school***** NEAR/2 counsel*) or (school* NEAR/2 facilitat*) or (teacher* NEAR/2 facilitat*) or (student* NEAR/2 facilitat*) or (pupil* NEAR/2 facilitat*) or (class* NEAR/2 facilitat*) or (school* NEAR/2 prevention) or (student* NEAR/2 mentor*) or (pupil* NEAR/2 mentor*) or (school* NEAR/2 activit*) or (afterschool NEAR/2 activit*) or (school* NEAR/2 program*) or (school* NEAR/2 therap*) or (student* NEAR/2 identif*) or (pupil* NEAR/2 identif*) or (teacher* NEAR/2 identif*) or (school* NEAR/2 deliver*) or (teach* NEAR/2 deliver*) or club or clubs) | (teacher* W/2 referral*) or (school* W/2 referral*) or (student* W/2 referral*) or (pupil* W/2 referral*) or (school* W/2 intervention*) or (preschool* W/2 intervention*) or (kindergarten* W/2 intervention*) or (class* W/2 intervention*) or "teach* intervention*" or "learning intervention*" or (student* W/2 support) or (pupil* W/2 support) or (school* W/2 support) or (preschool* W/2 support) or (kindergarten* W/2 support) or (lesson* W/2 support) or (class* W/2 support) or (teach* W/2 support) or (learning W/2 support) or (school* W/2 counsel*) or (school* W/2 facilitat*) or (teacher* W/2 facilitat*) or (student* W/2 facilitat*) or (pupil* W/2 facilitat*) or (class* W/2 facilitat*) or (school* W/2 prevention) or (student* W/2 mentor*) or (pupil* W/2 mentor*) or (school* W/2 activit*) or (afterschool W/2 activit*) or (school* W/2 program*) or (school* W/2 therap*) or (student* W/2 identif*) or (pupil* W/2 identif*) or (teacher* W/2 identif*) or (school* W/2 deliver*) or (teach* W/2 deliver*) or club or clubs |
|  | (school* adj3 referral*).ti,ab,kf. | (school* adj3 referral*).ti,ab,kf. | (school* adj3 referral*).ti,ab,id. | (school* adj3 referral*).ti,ab. | (school* adj3 referral*).ti,ab. |  |  |
|  | (student* adj3 referral*).ti,ab,kf. | (student* adj3 referral*).ti,ab,kf. | (student* adj3 referral*).ti,ab,id. | (student* adj3 referral*).ti,ab. | (student* adj3 referral*).ti,ab. |  |  |
|  | (pupil* adj3 referral*).ti,ab,kf. | (pupil* adj3 referral*).ti,ab,kf. | (pupil* adj3 referral*).ti,ab,id. | (pupil* adj3 referral*).ti,ab. | (pupil* adj3 referral*).ti,ab. |  |  |
|  | (school* adj3 intervention*).ti,ab,kf. | (school* adj3 intervention*).ti,ab,kf. | (school* adj3 intervention*).ti,ab,id. | (school* adj3 intervention*).ti,ab. | (school* adj3 intervention*).ti,ab. |  |  |
|  | (preschool* adj3 intervention*).ti,ab,kf. | (preschool* adj3 intervention*).ti,ab,kf. | (preschool* adj3 intervention*).ti,ab,id. | (preschool* adj3 intervention*).ti,ab. | (preschool* adj3 intervention*).ti,ab. |  |  |
|  | (kindergarten* adj3 intervention*).ti,ab,kf. | (kindergarten* adj3 intervention*).ti,ab,kf. | (kindergarten* adj3 intervention*).ti,ab,id. | (kindergarten* adj3 intervention*).ti,ab. | (kindergarten* adj3 intervention*).ti,ab. |  |  |
|  | (class* adj3 intervention*).ti,ab,kf. | (class* adj3 intervention*).ti,ab,kf. | (class* adj3 intervention*).ti,ab,id. | (class* adj3 intervention*).ti,ab. | (class* adj3 intervention*).ti,ab. |  |  |
|  | teach* intervention*.ti,ab,kf. | teach* intervention*.ti,ab,kf. | teach* intervention*.ti,ab,id. | teach* intervention*.ti,ab. | teach* intervention*.ti,ab. |  |  |
|  | learning intervention*.ti,ab,kf. | learning intervention*.ti,ab,kf. | learning intervention*.ti,ab,id. | learning intervention*.ti,ab. | learning intervention*.ti,ab. |  |  |
|  | (student* adj3 support).ti,ab,kf. | (student* adj3 support).ti,ab,kf. | (student* adj3 support).ti,ab,id. | (student* adj3 support).ti,ab. | (student* adj3 support).ti,ab. |  |  |
|  | (pupil* adj3 support).ti,ab,kf. | (pupil* adj3 support).ti,ab,kf. | (pupil* adj3 support).ti,ab,id. | (pupil* adj3 support).ti,ab. | (pupil* adj3 support).ti,ab. |  |  |
|  | (school* adj3 support).ti,ab,kf. | (school* adj3 support).ti,ab,kf. | (school* adj3 support).ti,ab,id. | (school* adj3 support).ti,ab. | (school* adj3 support).ti,ab. |  |  |
|  | (preschool* adj3 support).ti,ab,kf. | (preschool* adj3 support).ti,ab,kf. | (preschool* adj3 support).ti,ab,id. | (preschool* adj3 support).ti,ab. | (preschool* adj3 support).ti,ab. |  |  |
|  | (kindergarten* adj3 support).ti,ab,kf. | (kindergarten* adj3 support).ti,ab,kf. | (kindergarten* adj3 support).ti,ab,id. | (kindergarten* adj3 support).ti,ab. | (kindergarten* adj3 support).ti,ab. |  |  |
|  | (lesson* adj3 support).ti,ab,kf. | (lesson* adj3 support).ti,ab,kf. | (lesson* adj3 support).ti,ab,id. | (lesson* adj3 support).ti,ab. | (lesson* adj3 support).ti,ab. |  |  |
|  | (class* adj3 support).ti,ab,kf. | (class* adj3 support).ti,ab,kf. | (class* adj3 support).ti,ab,id. | (class* adj3 support).ti,ab. | (class* adj3 support).ti,ab. |  |  |
|  | (teach* adj3 support).ti,ab,kf. | (teach* adj3 support).ti,ab,kf. | (teach* adj3 support).ti,ab,id. | (teach* adj3 support).ti,ab. | (teach* adj3 support).ti,ab. |  |  |
|  | (learning adj3 support).ti,ab,kf. | (learning adj3 support).ti,ab,kf. | (learning adj3 support).ti,ab,id. | (learning adj3 support).ti,ab. | (learning adj3 support).ti,ab. |  |  |
|  | (school* adj3 counsel*).ti,ab,kf. | (school* adj3 counsel*).ti,ab,kf. | (school* adj3 counsel*).ti,ab,id. | (school* adj3 counsel*).ti,ab. | (school* adj3 counsel*).ti,ab. |  |  |
|  | (school* adj3 facilitat*).ti,ab,kf. | (school* adj3 facilitat*).ti,ab,kf. | (school* adj3 facilitat*).ti,ab,id. | (school* adj3 facilitat*).ti,ab. | (school* adj3 facilitat*).ti,ab. |  |  |
|  | (teacher* adj3 facilitat*).ti,ab,kf. | (teacher* adj3 facilitat*).ti,ab,kf. | (teacher* adj3 facilitat*).ti,ab,id. | (teacher* adj3 facilitat*).ti,ab. | (teacher* adj3 facilitat*).ti,ab. |  |  |
|  | (student* adj3 facilitat*).ti,ab,kf. | (student* adj3 facilitat*).ti,ab,kf. | (student* adj3 facilitat*).ti,ab,id. | (student* adj3 facilitat*).ti,ab. | (student* adj3 facilitat*).ti,ab. |  |  |
|  | (pupil* adj3 facilitat*).ti,ab,kf. | (pupil* adj3 facilitat*).ti,ab,kf. | (pupil* adj3 facilitat*).ti,ab,id. | (pupil* adj3 facilitat*).ti,ab. | (pupil* adj3 facilitat*).ti,ab. |  |  |
|  | (class* adj3 facilitat*).ti,ab,kf. | (class* adj3 facilitat*).ti,ab,kf. | (class* adj3 facilitat*).ti,ab,id. | (class* adj3 facilitat*).ti,ab. | (class* adj3 facilitat*).ti,ab. |  |  |
|  | (school* adj3 prevention).ti,ab,kf. | (school* adj3 prevention).ti,ab,kf. | (school* adj3 prevention).ti,ab,id. | (school* adj3 prevention).ti,ab. | (school* adj3 prevention).ti,ab. |  |  |
|  | (student* adj3 mentor*).ti,ab,kf. | (student* adj3 mentor*).ti,ab,kf. | (student* adj3 mentor*).ti,ab,id. | (student* adj3 mentor*).ti,ab. | (student* adj3 mentor*).ti,ab. |  |  |
|  | (pupil* adj3 mentor*).ti,ab,kf. | (pupil* adj3 mentor*).ti,ab,kf. | (pupil* adj3 mentor*).ti,ab,id. | (pupil* adj3 mentor*).ti,ab. | (pupil* adj3 mentor*).ti,ab. |  |  |
|  | (school* adj3 activit*).ti,ab,kf. | (school* adj3 activit*).ti,ab,kf. | (school* adj3 activit*).ti,ab,id. | (school* adj3 activit*).ti,ab. | (school* adj3 activit*).ti,ab. |  |  |
|  | (afterschool adj3 activit*).ti,ab,kf. | (afterschool adj3 activit*).ti,ab,kf. | (afterschool adj3 activit*).ti,ab,id. | (afterschool adj3 activit*).ti,ab. | (afterschool adj3 activit*).ti,ab. |  |  |
|  | (school* adj3 program*).ti,ab,kf. | (school* adj3 program*).ti,ab,kf. | (school* adj3 program*).ti,ab,id. | (school* adj3 program*).ti,ab. | (school* adj3 program*).ti,ab. |  |  |
|  | (school* adj3 therap*).ti,ab,kf. | (school* adj3 therap*).ti,ab,kf. | (school* adj3 therap*).ti,ab,id. | (school* adj3 therap*).ti,ab. | (school* adj3 therap*).ti,ab. |  |  |
|  | (student* adj3 identif*).ti,ab,kf. | (student* adj3 identif*).ti,ab,kf. | (student* adj3 identif*).ti,ab,id. | (student* adj3 identif*).ti,ab. | (student* adj3 identif*).ti,ab. |  |  |
|  | (pupil* adj3 identif*).ti,ab,kf. | (pupil* adj3 identif*).ti,ab,kf. | (pupil* adj3 identif*).ti,ab,id. | (pupil* adj3 identif*).ti,ab. | (pupil* adj3 identif*).ti,ab. |  |  |
|  | (teacher* adj3 identif*).ti,ab,kf. | (teacher* adj3 identif*).ti,ab,kf. | (teacher* adj3 identif*).ti,ab,id. | (teacher* adj3 identif*).ti,ab. | (teacher* adj3 identif*).ti,ab. |  |  |
|  | (school* adj3 deliver*).ti,ab,kf. | (school* adj3 deliver*).ti,ab,kf. | (school* adj3 deliver*).ti,ab,id. | (school* adj3 deliver*).ti,ab. | (school* adj3 deliver*).ti,ab. |  |  |
|  | (teach* adj3 deliver*).ti,ab,kf. | (teach* adj3 deliver*).ti,ab,kf. | (teach* adj3 deliver*).ti,ab,id. | (teach* adj3 deliver*).ti,ab. | (teach* adj3 deliver*).ti,ab. |  |  |
|  | club.ti,ab,kf. | club.ti,ab,kf. | club.ti,ab,id. | club.ti,ab. | club.ti,ab. |  |  |
|  | clubs.ti,ab,kf. | clubs.ti,ab,kf. | clubs.ti,ab,id. | clubs.ti,ab. | clubs.ti,ab. |  |  |
|  | *Subject headings:*  (Teaching/ or Learning/ or Students/ or School Teachers/ or Schools/ or Schools, Nursery/ or Education/) **and** (“Referral and Consultation”/ or Early Intervention, Educational/ or Internet-Based Intervention/ or exp Psychotherapy/ or Counseling/ or Primary Prevention/ or Secondary Prevention/) | *Subject headings:*  (teaching/ or learning/ or student/ or elementary student/ or middle school student/ or high school student/ or school child/ or school teacher/  school/ or kindergarten/ or primary school/ or middle school/ or high school/ or education/ or primary education/ or secondary education/) **and** (patient referral/ or early childhood intervention/ or web-based intervention/ or exp psychotherapy/ or counseling/ or prevention/ or primary prevention/ or secondary prevention/) | *Subject headings:*  (Teaching/ or Learning/ or Students/ or Elementary School Students/ or High School Students/ or Intermediate School Students/ or Junior High School Students/ or Kindergarten Students/ or Middle School Students/ or Preschool Students/ or Primary School Students/ or Teachers/ or Elementary School Teachers/ or High School Teachers/ or Junior High School Teachers/ or Middle School Teachers/ or Preschool Teachers/ or Special Education Teachers/ or Schools/ or Elementary Schools/ or High Schools/ or Junior High Schools/ or Kindergartens/ or Middle Schools/ or Nursery Schools/ or Education/ or Elementary Education/ or High School Education/ or Middle School Education/ or Secondary Education/ or Early Childhood Education/) **and** (Professional Referral/ or Self-Referral/ or exp School Based Intervention/ or exp Psychotherapy/ or exp Counseling/ or Prevention/) |  | *Subject headings:*  (teaching/ or learning/ or students/ or high school students/ or junior high school students/ or school children/ or teachers/ or kindergarten/ or exp schools/ or education/ or early childhood education/ or elementary education/ or preschool education/ or primary education/ or secondary education/)  **and**  (intervention/ or psychotherapy/ or therapy/ or counselling/ or prevention/) |  |  |

### S2. Forest Plot

Forest plot of effect sizes for all included studies (k = 38)

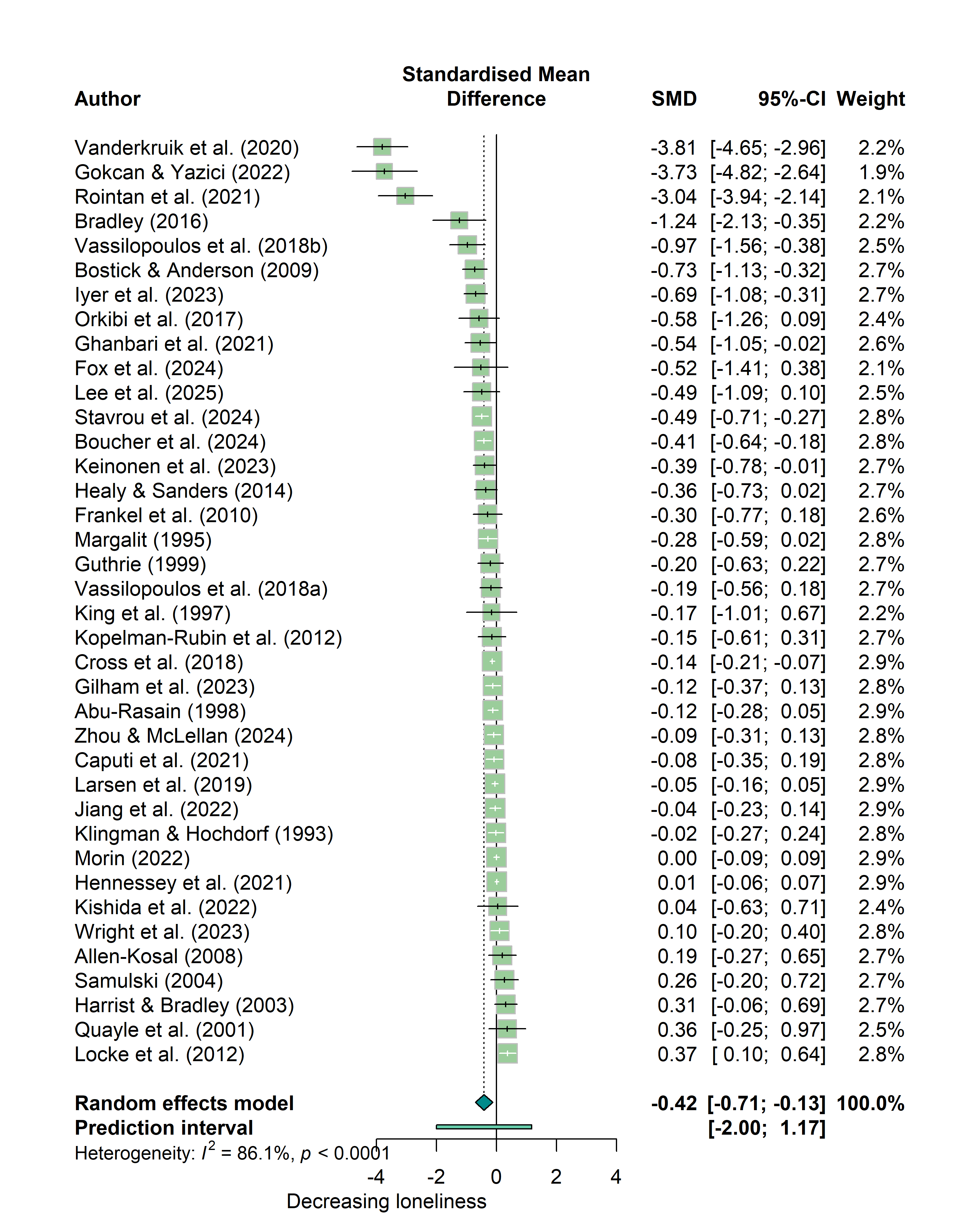

### S3. Funnel Plots

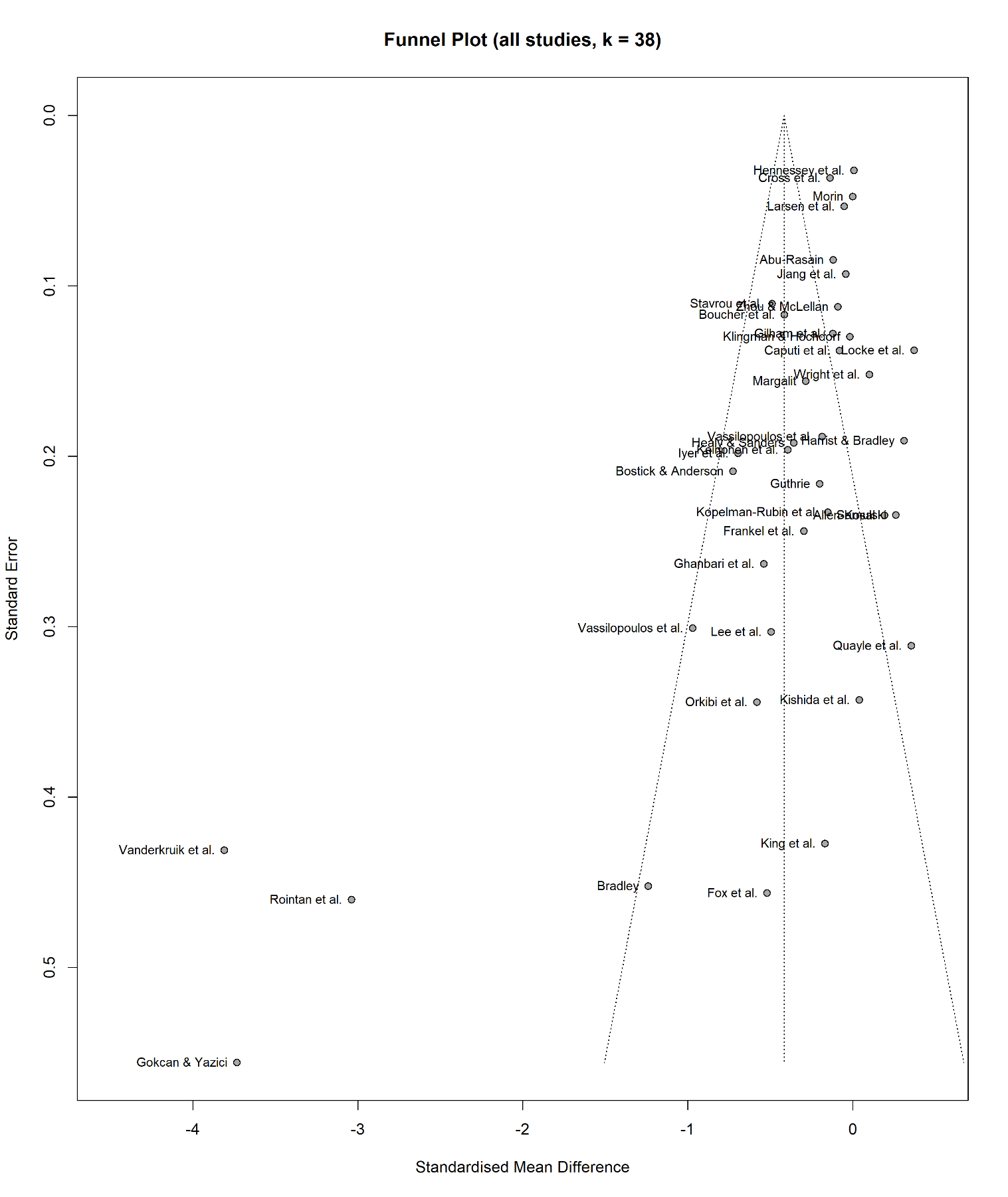

Note three studies with large standard errors on far left hand side of funnel plot, indicating three prospective small-study effects. Asymmetry also indicated by studies clustered to the right of the funnel with small SEs.

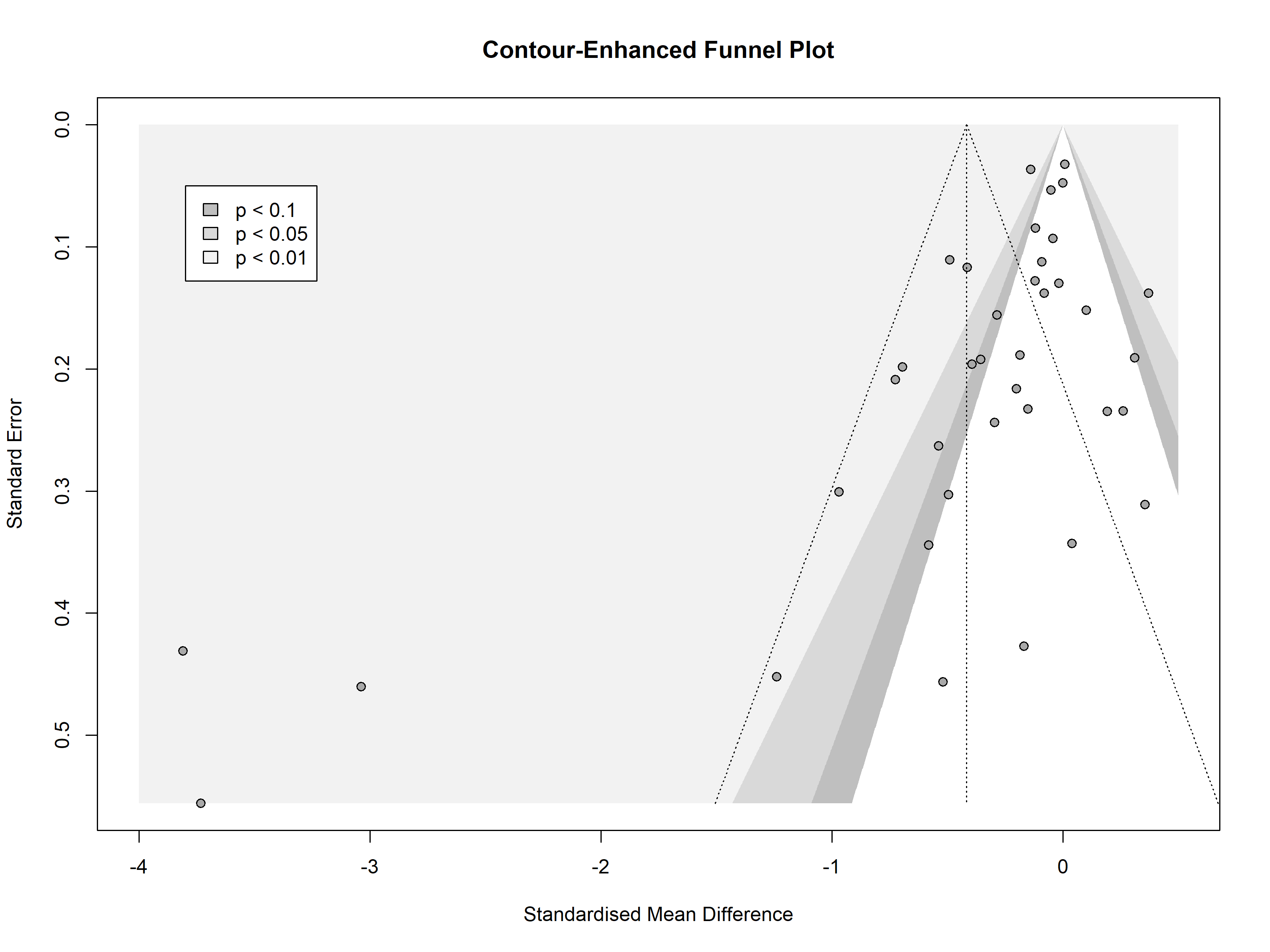

Contour-enhanced funnel plot illustrates how many of the studies are non-significant – suggests publication bias is not a major concern. Funnel plots assume effect size dispersion is due only to sampling error, but asymmetry can be produced by between-study heterogeneity; studies may be estimating different true effects, hence the use of a random-effects model.

### S4. Implementation Factors

Implementation factors extracted from included studies (*k* = 38)

| Authors (date) | Facilitators | Barriers |
| --- | --- | --- |
| Abu-Rasain (1998) | Peer delivery described as helpful, flexible and accessible by students | Lack of cooperation or enthusiasm from teachers |
| Allen-Kosal (2008) | None reported (NR) | Teachers unwilling to record data; could not address factors outside of school, e.g. family dynamics |
| Bostick and Anderson (2009) | Student self-referral option; parent and teacher involvement; evidence of impact in situ | Pressure on school counsellors to evidence impact on top of expectations out of role remit |
| Boucher et al. (2024) | Digital interventions private and cost-effective; parental support with help-seeking | Lack of time and resource in schools, especially in disadvantaged areas; fear of stigmatization |
| Bradley (2016) | Staff involvement and facilitation; experience of reciprocal social support; not feeling singled out | Interventions need to be right fit for school environment |
| Caputi, Cugnata & Brombin (2021) | NR | Short timeframe for training |
| Cross et al. (2018) | Active engagement of whole-school; students involved in developing content; tailoring content to each year level's specific needs | Lack of longer-term coaching and support for staff delivering intervention |
| Fox et al. (2024) | Flexible, non-intensive, relatively brief intervention | Virtual implementation provided no opportunity to practice behaviours; limited access to technology or private space |
| Frankel et al. (2010) | Active involvement of parents facilitated by scheduling; delivery in clinical setting cost effective | Narrow focus on high functioning children with ASD with limited transferability; short timeframe |
| Ghanbari et al. (2021) | Simple and inexpensive | NR |
| Gilham et al. (2023) | Facilitation by trusted school staff; offered as part of school curriculum in single-gender group to reduce possible stigma | NR |
| Gokcan and Yazici (2022) | Offers direct, pleasurable experience of social satisfaction | NR |
| Guthrie (1999) | Teacher involvement in developing intervention; delivery in naturalistic setting with social validity | Differential effects on students; differences in teaching style and temperament; short-term only |
| Harrist and Bradley (2003) | Positive peer behaviour reinforces intervention effect | Short-term delivery; insufficient opportunities for practice and reflection |
| Healy and Sanders (2014) | Significant parental involvement; no direct involvement of peers; informing teachers of bullying concern to prompt school action | Short time frame |
| Hennessey, Qualter and Humphrey (2021) | Equipped with general social skills to address feelings of loneliness | Limited insight to intervention fidelity and dose |
| Iyer et al. (2023) | Self-guided and virtual; accessible and convenient for age group | Reporting bias due to self-monitoring activity completion |
| Jiang et al. (2022) | NR | No integration or coordination with school staff or services; short-term and inflexible (manual-based) |
| Keinonen et al. (2023) | No resource required to identify at-risk young people; easy to disseminate and inexpensive | NR |
| King et al. (1997) | Groups provide positive social experience; close collaboration of parents, teachers, and peers | Withdrawing children from classrooms; limited opportunities to practice skills in natural setting |
| Kishida et al. (2022) | Schools encouraging teacher engagement and allocating time; teachers’ desire to gain knowledge | Lack of time and competing priorities |
| Klingman and Hochdorf (1993) | ‘Natural setting' of classroom; approachable delivery style | Short-term delivery |
| Kopelman-Rubin et al. (2012) | Use of manual allows consistency and flexibility; therapist tailors to individual’s unique needs | NR |
| Larsen et al. (2019) | Readiness for implementation; multi-tier approach targeting individuals as well as whole school | School transitions introduce many stressors; not enough time to train teachers and peer mentors |
| Lee et al. (2023) | Group-based delivery is social; availability of school support; easy and cost-effective programs | Limited support, personnel, and resource available from school |
| Locke, Rotheram-Fuller and Kasari (2012) | NR | NR |
| Margalit (1995) | NR | Students with different conditions and specific needs may require individualised interventions |
| Morin (2022) | Teachers' commitment to supporting positive climate; incorporation into schools' routines | Lack of fidelity assessment; short timeframe |
| Orkibi et al. (2017) | NR | Drama-based activities may provoke embarrassment or anxiety |
| Quayle et al. (2001) | Scheduling programme to promote student engagement; link content to curriculum learning outcomes | Floor effect of universal program; session conflicted with lunch-time activities |
| Rointan et al. (2021) | NR | NR |
| Samulski (2004) | Meeting teachers individually to review expectations; reinforcement systems for implementation | More intensive intervention to produce impact; teachers under pressure and resistant to change |
| Stavrou et al. (2024) | Person-centred facilitation style; age-appropriate activities; adapting to students' interests | Time and engagement demands challenging for schools; short timeframe |
| Vanderkruik, Gist and Dimidjian (2020) | "Near peer" delivery by college-aged women; cost-effective; facilitator engagement and adherence | Schedule coordination difficulties; students could not be guaranteed placement in a group with a friend |
| Vassilopoulos et al. (2018b) | Less directive facilitation style; practice strategies and make collective decisions in safe context | NR |
| Vassilopoulos, Brouzos and Koutsianou (2018a) | NR | Ceiling effect of universal intervention; peer preferences maintained between grades |
| Wright et al. (2023) | PPI and expert involvement; ease of incorporation into school schedule; pragmatic training and delivery | Limited personnel time for facilitator training and supervision; some LEGO sets too complicated |
| Zhou and McLellan (2024) | Pre-existing mental health course at school; easy to integrate without disruption | Meditation practices during lunch hour conflicted with naps and classwork |
